# A decade of Acceptability Research with Adolescents in Africa: Systematic review and evidence map

**DOI:** 10.1101/2021.06.22.21259328

**Authors:** Oluwaseyi Somefun, Marisa Casale, Genevieve Haupt Ronnie, Chris Desmond, Lucie Cluver, Lorraine Sherr

## Abstract

Interventions aimed at improving adolescent developmental outcomes are more likely to be successful if the young people they target find them acceptable. However, no standard definitions or indicators exist to assess acceptability, acceptability research with adolescents in LMICs is still limited, and no known reviews synthesise the evidence from Africa.

We conducted a systematic review of peer-reviewed studies assessing intervention acceptability with young adults (aged 10-24) in Africa, published between January 2010 and June 2020. This paper maps and qualitatively synthesizes the scope, characteristics, and findings of these studies, including definitions of acceptability, methods used, the type and objectives of interventions assessed, and overall findings on adolescent acceptability.

The review was carried out in line with the Preferred Reporting Items for Systematic Reviews and Meta-Analyses (PRISMA). Key word searches generated 4692 unique records and 55 final eligible studies, assessing 60 interventions. Most studies were conducted in Southern Africa, of which 32 jointly in South Africa and Uganda. The majority of interventions assessed for acceptability could be classified as HIV or HPV vaccine interventions (10), E-health (10), HIV testing interventions (8), support group interventions (7) and contraceptive interventions (6). The objectives of most interventions were linked to SDG3, specifically to HIV and sexual and reproductive health. Acceptability was overall high among these published studies. 22 studies provided reasons for acceptability or lack thereof, some specific to particular types of interventions and others common across intervention types.

Our review exposes considerable scope for future acceptability research and review work. This should include: extending acceptability research beyond the health (and particularly HIV) sector and to regions in Africa where this type of research is still scarce; including adolescents earlier, and potentially throughout the intervention process; further conceptualising the construct of acceptability among adolescents and beyond, and examining the relationship between acceptability and uptake.

**Key Questions:** *What is already known?:*

- Addressing the developmental needs of adolescents in African countries is critical if the continent is to achieve its sustainable development goals (SDGs).
- Many interventions aimed at strengthening adolescent developmental outcomes have not achieved desired impact, and adolescent involvement is often poorly envisaged and implemented.
- Uptake and effectiveness of interventions is likely to be higher if these interventions are acceptable to adolescent end-users.

*What are the new findings?:*

- Acceptability of interventions assessed in Africa was generally high among adolescents.
- Understanding of the intervention, ease of use, adequate emotional support, autonomy, confidentiality and protection from stigma were key overarching themes explaining why young people found interventions acceptable

*What do the new findings imply?:*

- Intervention developers and implementers across the continent should pay attention to these key aspects of interventions and their delivery.
- It is important to strengthen adolescents’ understanding of interventions, involve adolescents early on in intervention development, and engage with the broader context within which adolescent acceptability is shaped.
- There is a need for more acceptability research in important areas for adolescent development beyond (physical) health and, within the health sector, beyond HIV.

## Background

Addressing the developmental needs of adolescents in African countries is critical if the continent is to achieve its sustainable development goals (SDGs), and envisaged transformation articulated in the African Union’s overarching Agenda 2063 (1, 2). Adolescents make up the largest generation of their age group in history (3),and Sub-Saharan Africa (SSA) accounts for over 20% of the estimated 1.8□billion adolescents and young adults globally (4). Investing in adolescent wellbeing can have positive effects for individuals during adolescence and beyond, as well as potential positive societal effects. Interventions that reduce the consequences of poverty among adolescents, or lead to more positive behaviours, can influence development and wellbeing during adolescence and throughout the life course (5-7). Investment during adolescence can strengthen early childhood investments and reduce the burden of morbidity and mortality in adulthood (8). Moreover, it has been argued that investment in adolescents can help realize the ‘demographic dividend’ (9, 10), and reduce generational inequalities (11).

Substantial investment has been made globally in adolescent interventions focusing on areas such as sexual and reproductive health, nutrition, uptake of vaccines and prevention of substance abuse (12). Unfortunately these interventions have not always recorded impressive impact (13). Data from both high-income countries (HICs) and low- and middle-income countries (LMICs) reveal that many interventions focusing on adolescents are fragmented, poorly designed, and unequal in quality (14). One reason for this may be an insufficient understanding of the particular nature of adolescence (15).

Adolescence is a critical period characterised by rapid development of the physical, cognitive, social, and emotional capabilities that are instrumental across their life-course (3). Adolescence is also a time of gathering independence and the pathways to learning and experiencing such independence are varied, with experiential learning playing a key role. The rapid growth associated with this phase and its influences on behaviour need to be well understood in order to design timely and effective interventions (16).

Interventions may also fail to sufficiently consider the diverse environments in which adolescents live, that may shape their decisions and behaviour (17). This may lead to interveners missing important factors that, if unaddressed, will prevent the intervention from having the desired impact. Additionally, program implementers may lack the specialized skills necessary for delivering and sustaining these interventions (12). Adult interventions may not translate directly for adolescent audiences and programme adjustments may be inadequate.

Since most interventions seek to effect adolescent behavioural change, many of the obstacles to uptake and effectiveness could be addressed by affording sufficient importance to the perspectives and participation of adolescents themselves. When adolescents feel coerced to engage in a particular behaviour or accept interventions that they don’t identify with, they are more likely to resist the message of the proposed intervention, or to stop participating altogether (18). Instead, interventions that are acceptable to adolescent end-users are likely to have higher social validity (19), uptake and effectiveness (20, 21).

However, adolescent involvement and input in intervention design has been varied, and models of adolescent inclusion have been poorly envisaged and implemented. There is still a relatively low number of acceptability studies among adolescents in LMICs and specifically in Africa, particularly beyond the health sector (19, 20). To our knowledge no existing reviews comprehensively map the extant body of acceptability research in Africa and aggregate the evidence emerging from these studies. Furthermore, there is no clear and standard definition of acceptability (20) in Africa and beyond. This in turn raises several methodological challenges when setting out to assess acceptability, including the choice of measurement frameworks and tools (20). It also highlights the scope for further conceptualisation of this construct, particularly in specific populations and geographical regions.

We conducted a systematic review to identify studies that conducted primary research with adolescents and young adults (10-24) in Africa over the past decade (January 2010-June 2020), to assess the acceptability of interventions aimed at positively influencing their developmental outcomes. This paper maps and qualitatively synthesizes the scope, characteristics, and overall findings of studies identified. This includes evidence addressing the questions of whether and how the construct of acceptability is conceptualised and defined within these studies, the methods and indicators used, the type and key objectives of interventions assessed, as well as evidence on what adolescents find acceptable and why. Based on these findings, we aim to discuss implications for future adolescent-focused interventions in Africa and identify gaps for future acceptability research with this population.

## Methods

### Search strategy

The systematic review was carried out in line with the Preferred Reporting Items for Systematic Reviews and Meta-Analyses (PRISMA). We used the PICO (Population, Intervention, Comparison, Outcome) criteria (22) to help determine eligibility criteria for inclusion develop the search strategy and composite search terms developed (see Table S1). We searched 8 online databases (listed in Table S1), covering a wide range of behavioural science research, and searched the reference lists of eligible papers.

### Study selection and data extraction

Papers were selected based on the following inclusion criteria: if they (i) reported primary research assessing acceptability (based on the authors’ definition of the study or findings) of one or more intervention(s) with adolescents and young adults 10-24; (ii) assessed acceptability of intervention(s) aimed at positively influencing one or more development outcome(s), as defined by SDG indicators; (iii) reported on research conducted in Africa; (iv) were in the English Language; (v) were peer-reviewed and; (vi) were published between 1^st^ January 2010 and 30^th^ June 2020. We did not include limiters for study design or methodological tools, type of intervention or sector, or type of developmental outcome the intervention intended to influence. To be as inclusive as possible, we included studies that worked with broader samples (e.g., youth and adults) but disaggregated the results and reported findings specifically for the age group of interest (10-24).

We imported all references from the online databases into Endnote, where duplicates were identified and removed. Abstracts were reviewed independently by the two first authors to determine relevance. Full text of potentially eligible studies were retrieved and independently examined by the same two authors; areas of disagreement or lack of clarity were resolved through discussion by the two authors and – where necessary – the assessment of a third author. Reasons for exclusion of each paper not deemed eligible were recorded in an excel spread sheet. We developed a detailed extraction sheet, using Excel software, to extract key characteristics and findings of eligible papers. For reliability, the information for each paper was extracted separately by at least two of the first three authors and differences were resolved through discussion among the authors.

## Results

### Eligible studies included in the review

Figure 1 presents the PRISMA flow diagram describing the process of study selection and reasons for study exclusion. A total of 4692 titles and abstracts were screened after removing duplicates, 278 articles were subjected to a full-text review, and a final 55 studies were considered eligible for inclusion in the review.

**Figure 1:**
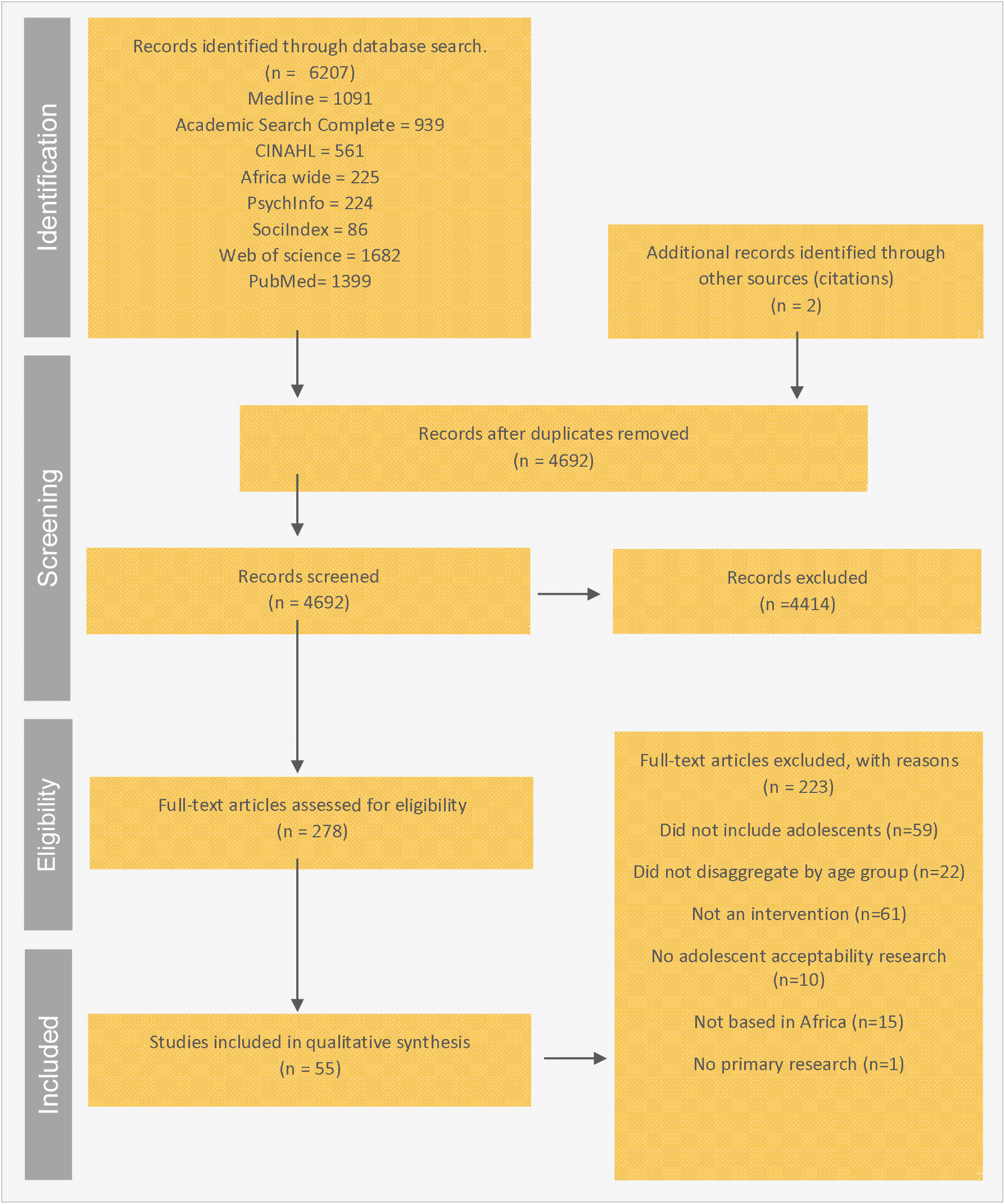
The PRISMA flow diagram describing the process of study selection.

### Study characteristics: publication year, location and sample

Below we present a summary of key characteristics of the 55 eligible studies included in our review. More than half of the papers were published between 2018-2020 with 22% of the papers published in 2019, as shown in the supplementary figure S1.

Fig.2 below provides a visual representation of the location of studies on the continent. There is a clear concentration of acceptability studies in South and East Africa, with approximately half of identified studies conducted in South Africa (19) and Uganda (13). Only seven studies were from West and Central Africa and only one from North Africa.

**Figure 2:**
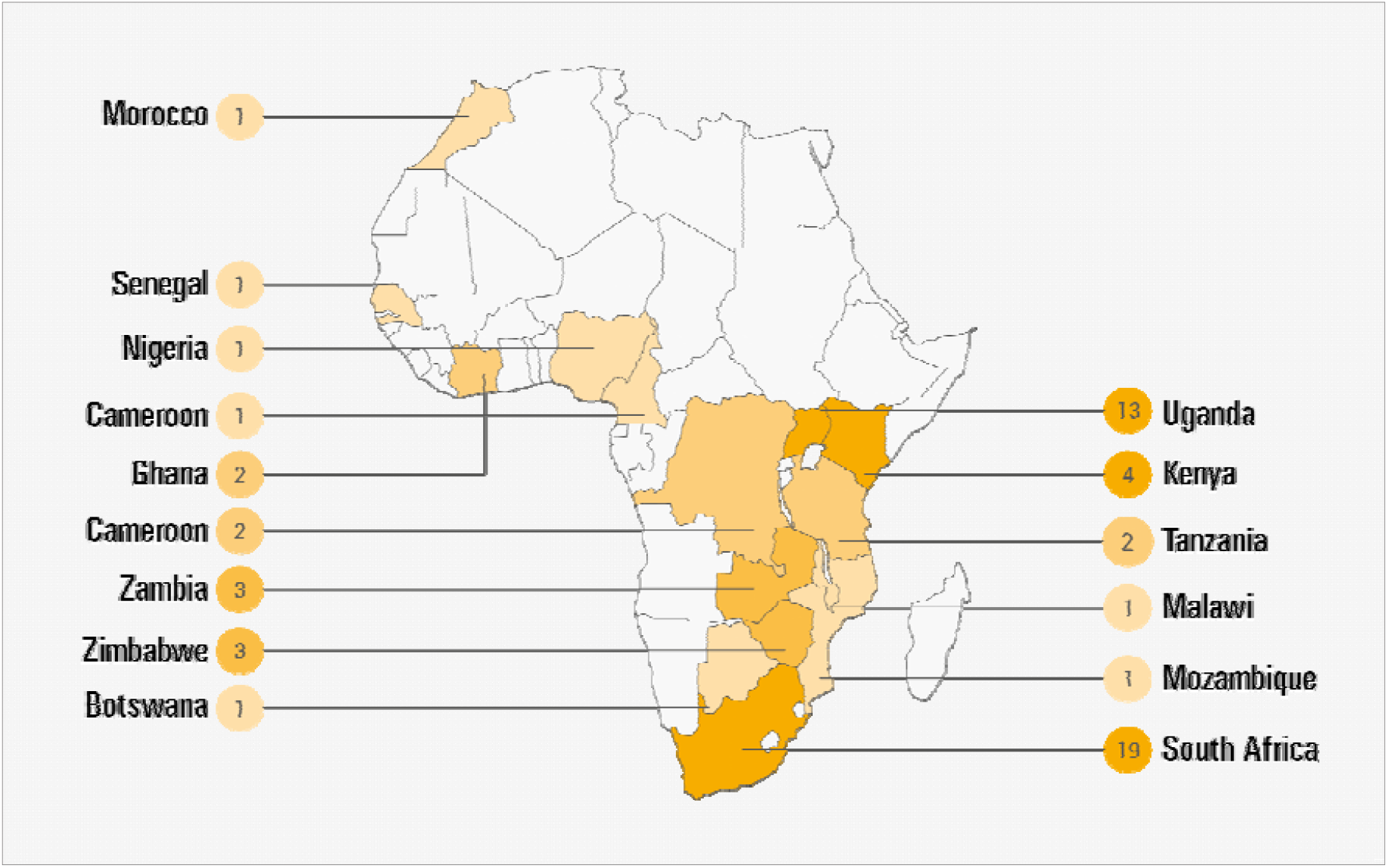
Study Location.

The supplementary table S2 provides information on study characteristics and overall findings for the entire list of eligible studies, and by each type of intervention category (as indicated below) in separate sheets. Most (41) study samples included male and female participants, while 11 studies worked only with females and three with males only. 44 studies worked with samples that fell entirely within the specified age range (10-24), while 11 included studies worked with broader samples (e.g., youth and adults) but disaggregated the results and reported findings specifically for the age group of interest. To be as inclusive as possible, we included 10 studies that did not clearly specify the exact age range of participants, but for which available information indicated that the sample would have been entirely or almost entirely within this range (e.g. secondary school and university students (23-28) 0r where sample descriptive data indicated a sample consisting almost entirely of participants 24 or younger (29-31).

While our inclusion criteria focused on primary acceptability research with adolescents and young adults, it should be noted that 25 studies also collected acceptability data from other stakeholders. These include caregivers or other family members (32-40), teachers, facilitators (26, 41, 42), community leaders or gate keepers, (28, 43), peer mentors, service providers and healthcare workers (29, 44-51).

### Types and objectives of interventions assessed for acceptability

We categorised interventions assessed for acceptability both by type of intervention, based on their key components (see Figure 3), and stated objectives of the interventions (see Figure 4). In terms of type of intervention, interventions were classified as HIV or HPV vaccine interventions (10), E-health (10), HIV testing interventions (8), support group interventions (7), contraceptive interventions (6), voluntary medical male circumcision programs (VMMC) (4), school-based sexual and reproductive health education (4), economic support programs (4) and pre-exposure prophylaxis (PrEP) (2). Five studies did not fit into the above intervention categories and were grouped as ‘other’; they consisted respectively of nutritional therapy, a psychosocial - home based care intervention, a counselling support intervention to address substance abuse, cervical cancer screening and a rectal microbicide intervention for HIV prevention. It should be noted that two of the studies reviewed assessed more than one intervention (45, 52) (3 and 4 respectively), so that the total number of interventions assessed for acceptability was 60.

**Figure 3:**
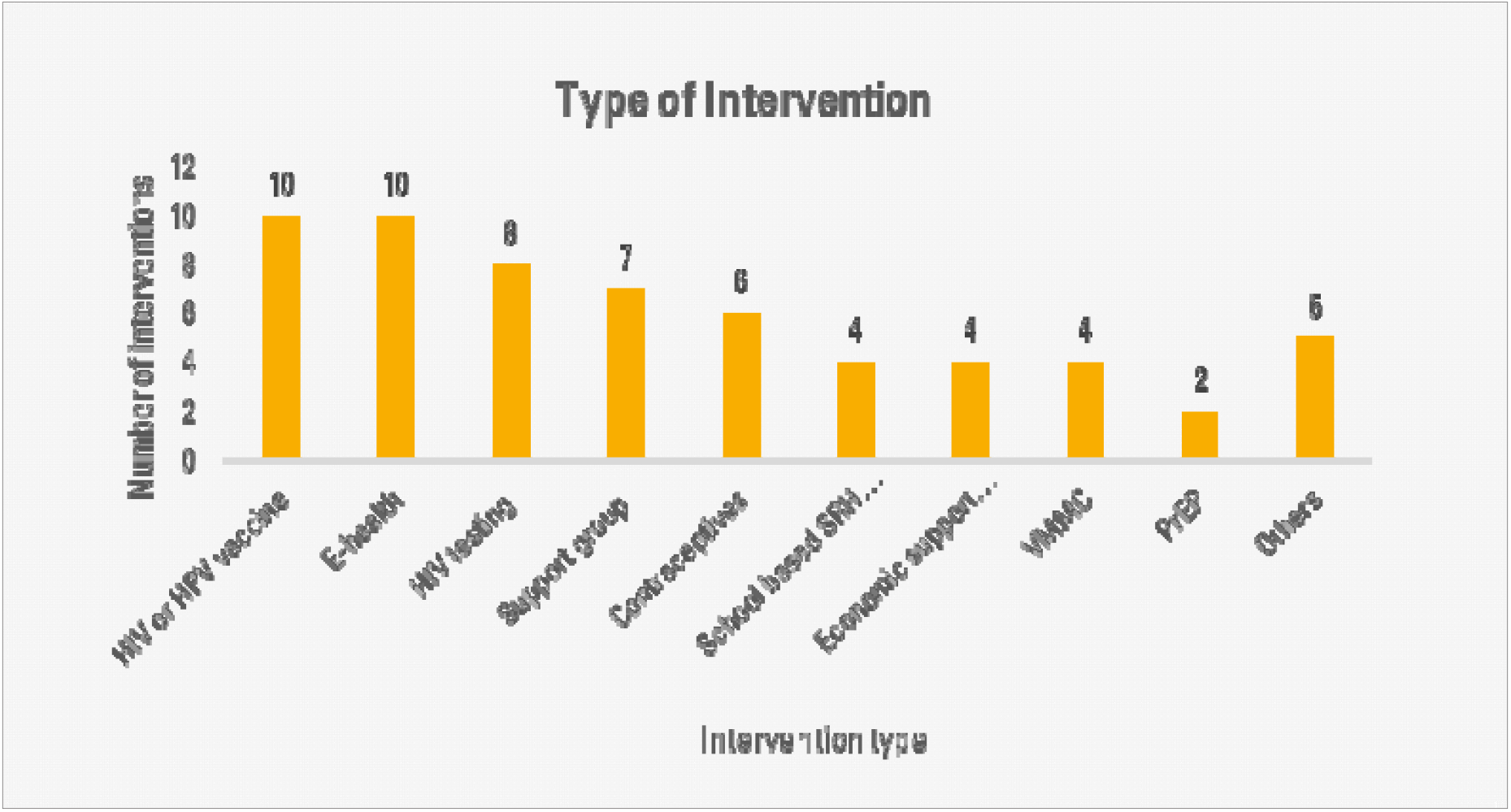
Intervention Types.

**Figure 4:**
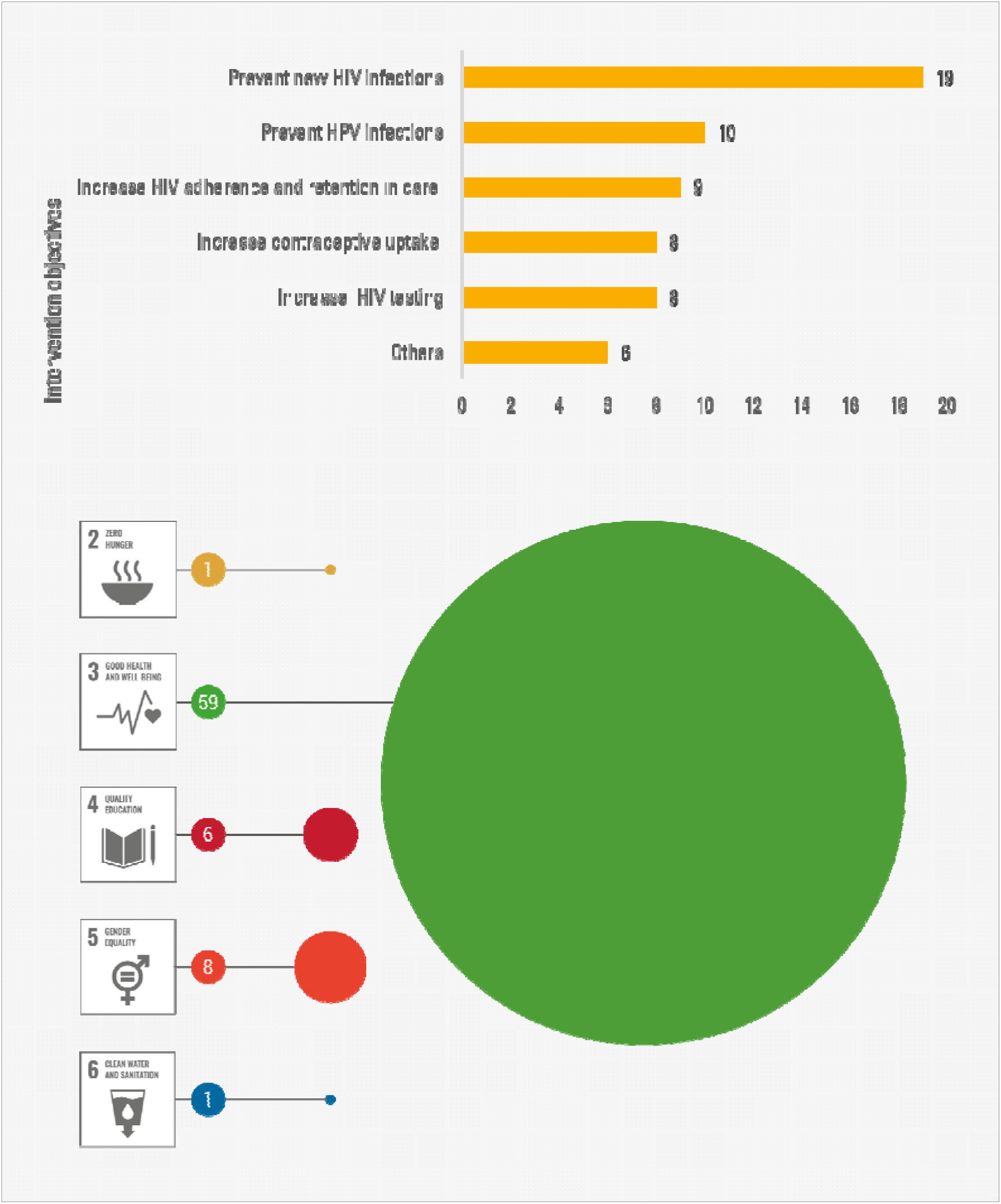
Intervention objectives and number of interventions linked to each SDG.

More detail on intervention sub-types is included in Table S2. For example, E-health interventions included game based (1), SMS based (7) and internet-based (2) programs.

All 7 support group interventions provided psychosocial or educational support related to HIV, and 5 worked only with young adults living with HIV. One group intervention was delivered through both a social media platform and in-person meetings (53), one was a family based support intervention with adolescent-parent dyads (33), four were linked to public healthcare facilities (42, 47, 54, 55) and one was a community intervention (43).

The primary objectives of most interventions were focused on HIV- or sexual and reproductive health-related outcomes (see Figure 4): 19 primarily aimed to prevent new HIV infections, ten to prevent HPV infection, nine to increase HIV treatment adherence and retention in care, eight to increase the uptake of HIV testing, eight aimed at increasing contraceptive uptake and reducing early childbearing and six provided psychosocial support for adolescents living with HIV (42).

The objectives of almost all interventions were therefore linked to indicators within SDG3 (ensuring healthy lives and promoting well-being). However, one study could also be linked to SDG2 (food security and improved nutrition), 6 to SDG4 (inclusive and equitable quality education), 8 to SDG5 (gender equality) and 1 to SDG6 (access to water and sanitation).

### Definitions and conceptual frameworks for acceptability

Only seven of the 55 reviewed studies provided an explicit definition of acceptability and only six used a conceptual framework (as indicated in Table S2). Three definitions focused on the preference for or willingness to use the intervention: Tonen-Wolyec et al (2019) defined acceptability as consenting to and using the (HIV self-testing) intervention; Smith, Wallace (30) defined it as the preference for using the (HIV self-testing) device ^33^; and Katahoire et al (2013) defined acceptability as the willingness or reluctance to use and complete the intervention (in this case the 3 doses of HPV vaccine) (56).

Two definitions focused mainly on responses to the intervention. MacCarthy et al (2020) (48) referred to a definition and framework developed by Sekhon et al (2017)(20) and defined acceptability as the cognitive and emotional responses to an intervention (20, 48). Parker et al (2013) (42) defined acceptability as how the intended individual recipients react to a program, guided by the Bowen feasibility framework (57). A further two studies conceptualized acceptability as an implementation outcome and focused on value, appeal and likeability: Kibel et al (2019)(58) referred to the perception among stakeholders that a certain element of the program was valued, agreeable, or satisfactory, while Sabben et al (2019)(34) defined acceptability as appeal, relevance, value, usability, and understandability, based on the Technology Acceptance Model’s (TAM) framework (59).

Three studies referred to a conceptual framework but did not provide an explicit definition of acceptability. In their assessment of individual and environmental barriers and facilitators related to use of a school-based contraception clinic, Khoza et al (2019) referred to the social ecological framework (60). Sayles et al’s (2010) study was guided by value-expectancy and social marketing theories (61); the authors investigated vaccine attitudes, normative vaccine beliefs, and perceived risk and severity of HIV as determinants of HIV vaccine uptake. Turiho et al’s (2017) study used the symbolic interactionism theory (62) and some aspects of the Health Beliefs Model (HBM) to explain how community members’ perceptions and their interaction shape vaccine acceptability.

### Study design, methods and indicators

Sixteen studies included in this review (29%) assessed ‘anticipated’ or prospective acceptability among adolescents who had not (yet) received the intervention (20). 18 studies (33%) assessed acceptability concurrently, during the delivery of the intervention, while 14 (25%) assessed acceptability post-intervention, retrospectively. The remaining seven (13%) of the studies assessed interventions prospectively and retrospectively; among these, two studies worked with separate groups of adolescents who had received and not yet received the intervention (52, 63), while the remaining 5 interviewed adolescents at two different stages of the intervention (40, 44, 55, 64, 65). Five studies involved adolescents in the study design (43, 50, 53, 55, 65).

20 studies described their methodology as solely qualitative, 18 as quantitative and 17 as mixed methods. 11 of the qualitative studies used only focus group discussions (FGDS), 7 used only in-depth interviews (IDIs) and 2 used both methods. Most of the quantitative studies (15) employed structured survey questionnaires. The mixed methods studies combined FGDs or IDIs with survey questionnaires, online surveys and evaluation reports.

As detailed in the supplementary table S2, a wide range of questions and indicators were used to measure acceptability. None of the studies used a standardized previously validated instrument, although two papers drew from existing instruments (66, 67). The majority of questions asked across studies covered participants’ overall perceptions and experience of the intervention, willingness to use the intervention, understanding of the intervention, barriers and facilitators of access and use, the perceived effectiveness of the intervention and willingness to recommend or distribute it to others.

### Acceptability findings

Overall, acceptability of interventions assessed was high. Of the 55 studies, 30 assessed acceptability quantitatively and reported on the proportion of young adults in the sample that found the intervention acceptable. While some studies quantified acceptability through a single percentage, based on one question or indicator, a number of studies reported a range, based on multiple questions or indicators. One of the reviewed studies reported 100% acceptability (33), while acceptability ranged from 64% - 100% in 25 studies and 46% - 61% in 2 studies (27, 52, 68, 69). Only two studies clearly reported acceptability below 50%: at 37% for a contraceptive intervention in Tanzania (70) and 27% for an HPV vaccine study in Morocco (71). Reasons given for low acceptability of the contraceptive intervention were that adolescents and their peers were too young to be sensitized about condoms, that condoms would not be used properly and that using contraception was a sin (70). Reasons were not provided by adolescents for the Moroccan study; however, in quantitative analysis, older age, female gender, studying at a public (versus private) school and lower educational attainment were associated with lower odds of acceptability for the HPV vaccine (71).

The remaining 25 studies did not quantify acceptability. However, the authors of two of these studies reported that adolescents found the interventions to be unacceptable, based on their overall findings. One study in South Africa assessed contraceptive interventions (32); a key reason for low acceptability was the belief that a school-based contraceptive clinic (SBCC) could promote promiscuity by sending a message that ‘teenage sex was acceptable’ and making contraceptives easily accessible (32). The second study assessed a psychosocial home based care intervention in Tanzania (72), which adolescent participants felt did not align well with their expectations. They believed the intervention to be more relevant to their caregivers and were disappointed in the lack of financial support in a context of widespread poverty (72).

Findings of the remaining 51 studies overall indicated high levels of acceptability. Some of these studies also provided various reasons as to why adolescents found the interventions acceptable (n=22) or (for a minority of adolescents) not acceptable (n=20). These are presented in Table 1, by type of intervention, for studies with both low and high overall acceptability. The main reasons e-Health interventions were acceptable to adolescents were: knowledge gained from the intervention regarding their sexual health (34, 65), the privacy these interventions provided (23, 48) and knowing how to make use of the intervention (25, 34). Adolescents who instead did not find these interventions acceptable felt that the content was not culturally appropriate (23, 25, 65), highlighted technological glitches (48, 50, 65) or were concerned with inclusiveness where, for example, not all the young adults had access to a necessary device or risked unintended disclosure of private information when sharing devices (65, 73).

**Table 1:**
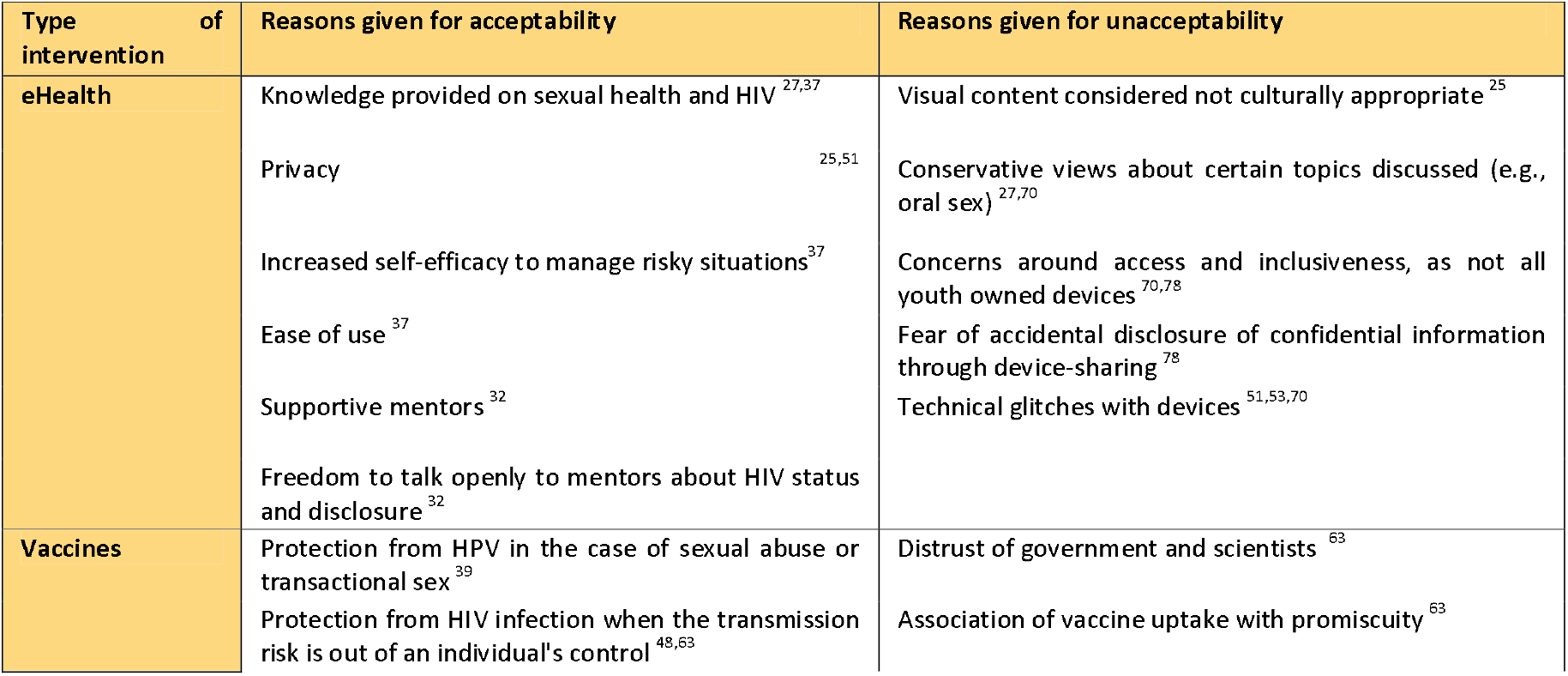
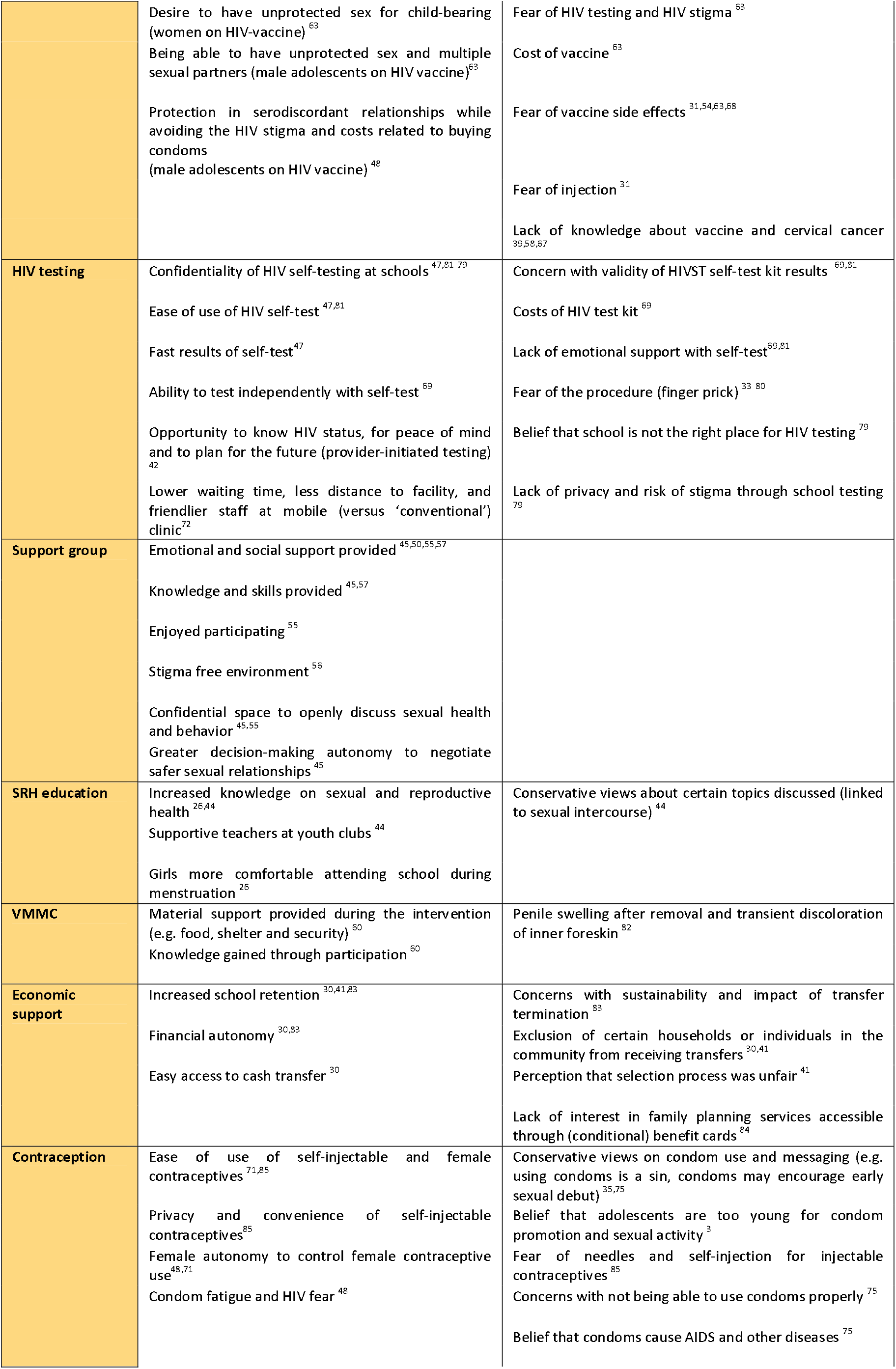
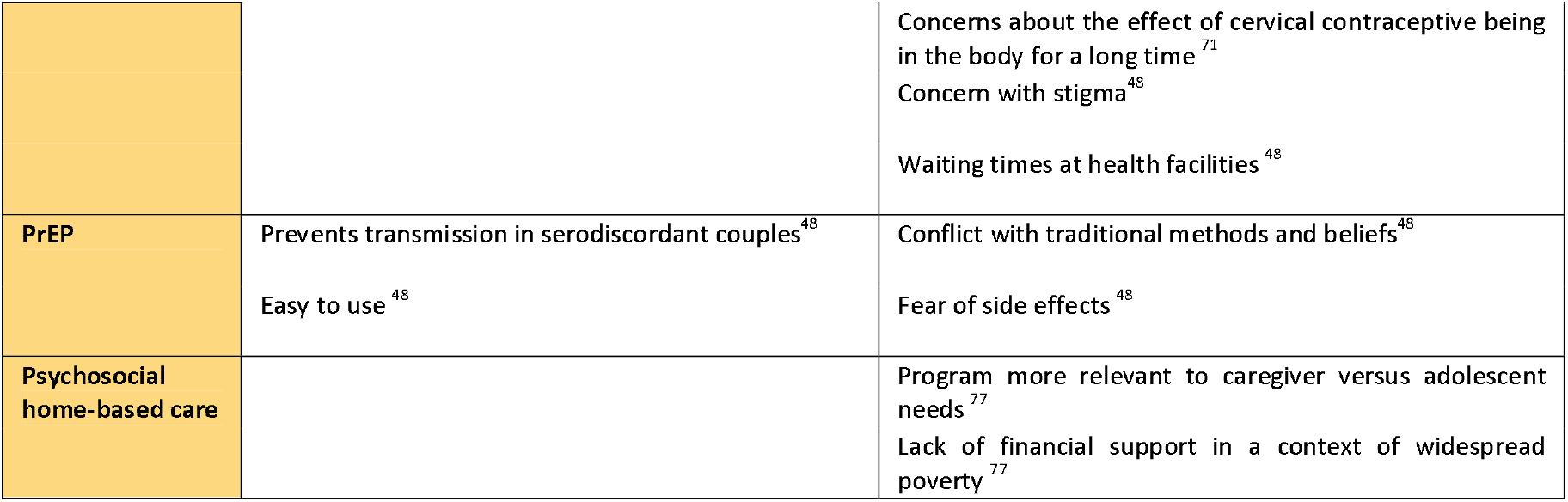
Reasons provided by adolescents for acceptability and unacceptability of interventions, by type of intervention.

Confidentiality, appropriateness, privacy and decision-making autonomy were among the reasons adolescents found HIV testing interventions (including self-testing and testing in schools) acceptable (42, 44, 53, 64, 74). Fear of the procedure, concerns with the cost and validity of the test, and inadequate emotional support were reasons given for lack of acceptability (64, 75, 76). Support group interventions were considered acceptable because of the emotional support provided and because young adults found the groups to be empowering and were able to discuss HIV-related issues in a stigma-free environment (42, 47, 53, 55).

Knowledge was a key reason for high vaccine acceptability for both HPV and HIV vaccine interventions. For example, adolescents’ understanding that HPV vaccines could prevent cervical cancer and HIV made them more likely to accept the interventions (63). Conversely, lack of knowledge or understanding of the intervention was linked to low acceptability (36, 52, 56). Other reasons given for acceptability were greater female autonomy and agency to protect themselves, in the event of sexual violence or transactional sex, and encouragement of peers (36, 58, 63). On the other hand, perceived cost, myths and distrust of vaccine providers, and fear of side effects, were themes raised to explain low acceptability (61, 77).

Reasons for acceptability of economic support interventions included financial autonomy (78) and the freedom to decide how to use cash transfers (28). However, concerns around the process of selecting which individuals or households were to receive transfers, as well as inclusion, sustainability and effects on social relations and social equity within the community (38, 78), were factors that threatened acceptability.

## Discussion

Findings of this review indicate two positive trends. The first is an increase, over the past decade, in the number of acceptability studies with adolescents on the continent. Though numbers are overall low, this could signal increasing recognition of the value of engaging young people when designing and implementing interventions intended for them. The second is that acceptability of interventions assessed was generally high. This suggests an overall good alignment of interventions with adolescent needs and preferences. However, we should also be aware of the possibility of publication bias (79, 80), as research showing less favourable acceptability results may be less likely to be written up and published. A key limitation of this review is that we did not include grey literature, given available resources, the review’s already broad scope, and to ensure a minimum quality of studies included. We also did not conduct a quality assessment, given the heterogeneity of interventions assessed and study designs; however, we note that this is not a requirement of a mapping review, which aims to summarise available evidence in an area versus focus on a particular research question (81-83).

### Acceptability findings

Despite the diversity of intervention settings, types of interventions and modes of delivery across studies, several common themes emerged from reasons given by adolescents to explain why specific interventions were acceptable to them. These included the product or intervention being easy to use, knowledge of the intervention or knowledge provided by the intervention, the intervention allowing for (greater) autonomy, adolescents feeling supported while participating in the intervention and feeling assured that their privacy and confidential information would be protected. Although reasons for ‘unacceptability’ were more diverse, overarching themes could also be identified among these, for example: conservative views about the intervention or its content; concerns around intervention costs, access and inclusiveness; fear of pain and side effects (for biomedical interventions); stigma, myths or distrust; and lack of knowledge or support. While certain drivers of unacceptability mirrored those of acceptability (e.g. knowledge and support), these drivers mostly differed, suggesting that acceptability and unacceptability are not necessarily represented by one continuum.

These findings suggest that intervention developers and implementers across the continent should pay attention to key aspects of interventions and their delivery that adolescents clearly care about, and seek to address these from the intervention development phase. They should ensure that adolescents are provided with adequate knowledge, training and resources to properly understand the intervention and feel confident in their ability to use it, that they have access to sufficient logistical and emotional support while participating, and that their confidential information is protected, so that they are in turn protected from much-feared stigma and other potential negative social consequences. Moreover, they should bear in mind that adolescents value autonomy and that this has a gender dimension. Autonomy relates not only to being able to choose to participate in and use an intervention, but also being empowered by the knowledge it may provide and the greater control it may afford young people (particularly young women) in managing high risk situations and unequal relationships.

It may also be worth paying particular attention to acceptability findings for specific types of interventions, given current African and global public health challenges. For example, the role of digital technology in achieving many of the SDGs is well documented (84) and merits particular attention in the context of the Covid-19 pandemic (85, 86). While young people remain the most connected population group to digital platforms(87), there is a clear digital divide, as more than 60% of young adults in Africa do not have access to internet (88, 89). Findings of this review show overall high acceptability of e-Health interventions (34, 50), as adolescents highlighted opportunities presented by digital technology, for example by reducing the cost of in-person interaction (53). Yet concerns raised around connectivity issues, lack of access to devices and unintended disclosure of confidential information (53, 73) represent challenges for the acceptability, equitable access and effectiveness of e-Health programs. It is therefore important for intervention providers to assess these challenges early on, and to explore ways of potentially increasing access to devices or technologies within the intervention itself or by supporting concurrent initiatives (65).

Low acceptability of several interventions aimed at increasing contraceptive use and HIV testing also merits particular attention, since HIV transmission and relatively low rates of HIV testing and linkage to antiretroviral therapy (ART) remain a concern among young adults (90, 91). Several studies included in this review highlighted, for example, adolescents’ fear of stigma and lack of privacy regarding HIV testing interventions in schools (74), concerns about not being able to properly perform oral HIV testing on their own (76) and conservative views of contraceptive promotion and use (32, 70). These perspectives are likely shaped by inadequate understanding of interventions, but also by social norms surrounding sexuality and contraception within adolescents’ homes, schools and communities (92, 93). Also, fear of vaccines and their side effects (94, 95) are important to note and address, in relation not only to HPV prevention, but also to the current Covid-19 vaccine rollout.

All of the above examples highlight the importance of strengthening adolescents’ knowledge of interventions and how to interact with them, but also of understanding and engaging with the broader context within which adolescent acceptability is shaped (92). One way to achieve this is to involve adolescents (preferably potential end-users) early in the design and planning phase of the intervention and – if possible - at various stages of the intervention life cycle. Yet, as indicated above, less than half of the studies in this review (42%) assessed prospective acceptability and very few studies involved adolescents in the study design and/or at multiple phases of the intervention. There is clearly potential to allow for more meaningful and consistent adolescent engagement, if young people are to have a stronger role in shaping the development, adaptation and scale up of interventions (20).

A second key approach would be to engage early on and assess acceptability with other stakeholders who are central to an intervention being well-targeted, well-implemented and accepted by adolescents and the broader community. These may include intervention implementers and facilitators, but also caregivers, partners and peers, teachers and community leaders. As noted above, 25 studies in this review also assessed acceptability of other types of stakeholders. Future review analyses and acceptability studies could further focus on acceptability among these groups of individuals, and its implications for adolescent acceptability and intervention success.

### Gaps and key areas for future research

Our review highlights several key gaps and related areas for future intervention acceptability research. First, there appears to be a gap in geographical coverage, particularly in West, Central and North Africa. However, we note that confining our search to English language publications may have excluded some studies from African countries where French is the first language. Given that adolescent needs and preferences are likely to differ across areas with very different social and cultural norms and faith contexts (96), we cannot simply extrapolate acceptability findings to other countries or communities across the continent.

Second, there is clearly scope for more acceptability research in important areas for adolescent development beyond (physical) health and, within the health sector, beyond HIV. As important as reducing HIV transmission and increasing testing and treatment adherence may be in this population (90, 91), they are clearly not the only dimensions of adolescent health and broader wellbeing that merit attention and investment. There is a glaring lack of acceptability studies in areas of adolescent development beyond SDG 3. These include education access and outcomes, employment opportunities, access to water and other services, gender equality and protection from violence, social protection and mental health (97).

The focus on specific types of interventions likely reflects, to a large extent, global health funding and research priorities over the past decades. There has been a considerable amount of international aid dedicated to addressing HIV (98, 99) and particular concern around the acceptability of HIV interventions. Moreover, the concentration of acceptability research in specific countries in Africa is likely a reflection of disparities in independent research infrastructure and capacity across the continent (100, 101). It would also seem that ‘acceptability’ is a concept and term that has gained traction primarily within the health sector (20). The extension of acceptability research to geographical and developmental areas where it is currently scarce therefore cannot be addressed solely by decisions of individual research teams, but will to some extent require a change in global health and funding priorities, and the ‘adoption’ of acceptability research by other sectors.

A third gap highlighted by this review is the considerable scope to further conceptualise the construct of acceptability, by more clearly defining it and identifying its key components. Our review reinforced the absence of a clear or standard definition of acceptability, or common tools and indicators. In fact, the large majority of papers included in this review (48) referred to the concept of acceptability without defining it at all, requiring the reader to review the questions and indicators used to gain some understanding of how the construct of acceptability was conceptualised and operationalized. As highlighted by other authors, this lack of common definitions and frameworks makes the selection of measurement indicators for empirical enquiry in this area more difficult and the comparability of acceptability results challenging (102, 103). There have been recent efforts to address these gaps; in particular, Sekhon and colleagues’ theoretical framework for acceptability (TFA), published in 2017 (20), has made a valuable contribution to the scarce conceptual literature in the field. However, there is still much work to be done to apply and test the framework in specific populations. For example, its relevance and completeness in investigating acceptability among adolescents, in less-resourced settings and beyond the (biomedical) health sector is still unclear. Also unclear is the important link between intervention acceptability and uptake, considering that willingness to use the intervention is often included among questions used to assess acceptability (see table S2). Lastly, it is encouraging to note that a relatively large number of studies in our review used mixed methods approaches to assess acceptability; however, there is clearly still scope to employ and combine more innovative methodologies (55, 65).

## Conclusion

As the first systematic review to aggregate and synthesise a decade of acceptability studies with adolescents in Africa, we believe this study makes a valuable contribution to the African and global literature on acceptability. It highlights the overall high level of acceptability of the interventions assessed, and some of the reasons why adolescents and young adults may or may not find interventions acceptable– both specific to particular types of interventions and common across intervention types.

However, it also exposes considerable scope for future acceptability research and review work, to extend and strengthen the existing body of evidence. This should include: extending acceptability research beyond the health (and particularly HIV) sector and to countries in Africa where this type of research is still scarce; including adolescents and other potential key stakeholders earlier, and potentially throughout, the intervention process; further conceptualising the construct of acceptability; and investigating the relationship between acceptability and intervention uptake and success.

## Data Availability

NA

## Acknowledgements

ODS, MC, GH, CD, LC and LS receive funding from the UKRI GCRF Accelerating Achievement for Africa’s Adolescents (Accelerate) Hub (Grant Ref: ES/S008101/1).

## Contributors

ODS and MC were involved in the conceptualisation of the paper, the data extraction and analysis, and the manuscript writing; GH was part of the data extraction and analysis, and reviewed and revised the manuscript; CD reviewed and revised the manuscript; LC reviewed and revised the manuscript; and LS reviewed and revised the manuscript. All authors read, reviewed and approved the final manuscript.

---

**Table S1.**
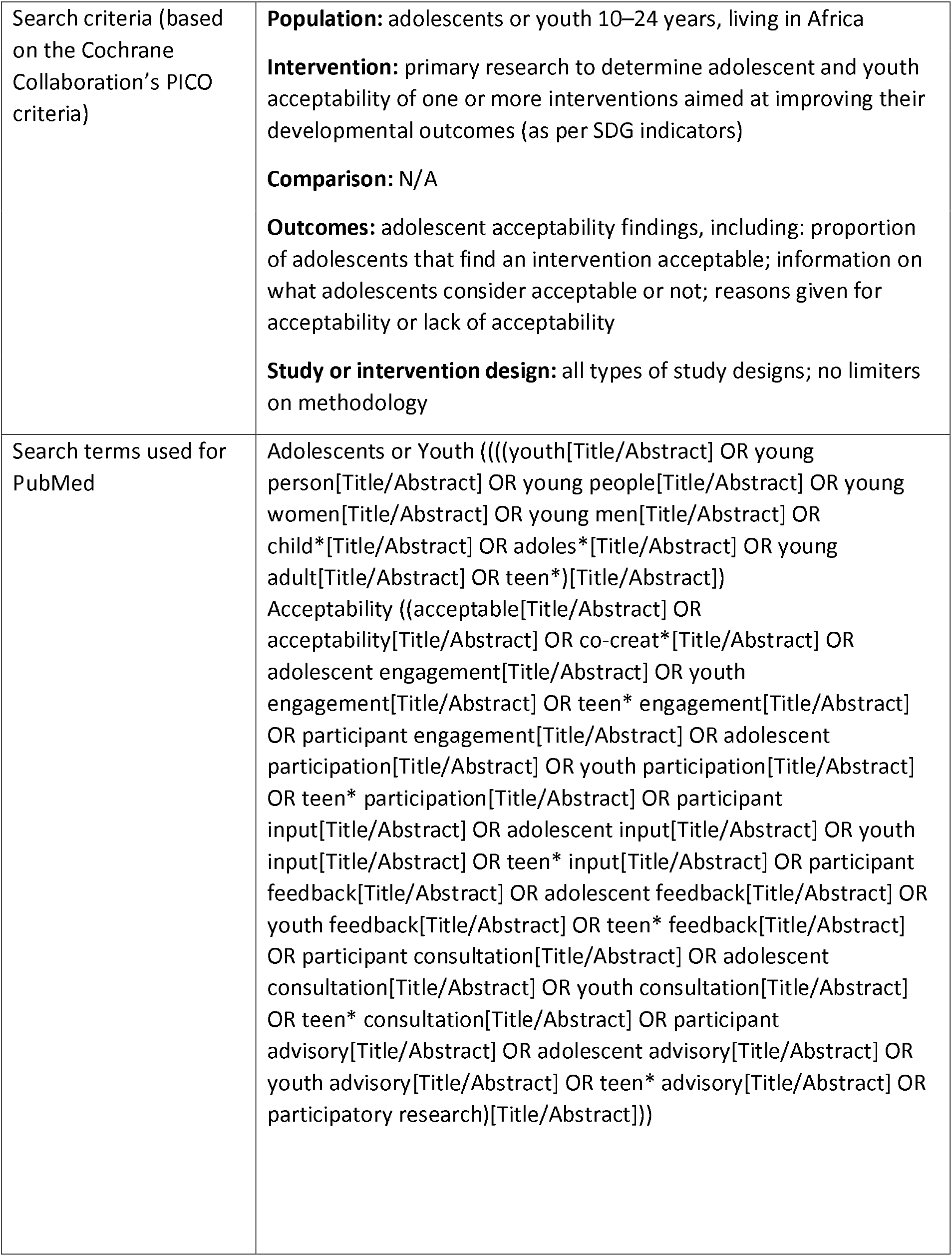
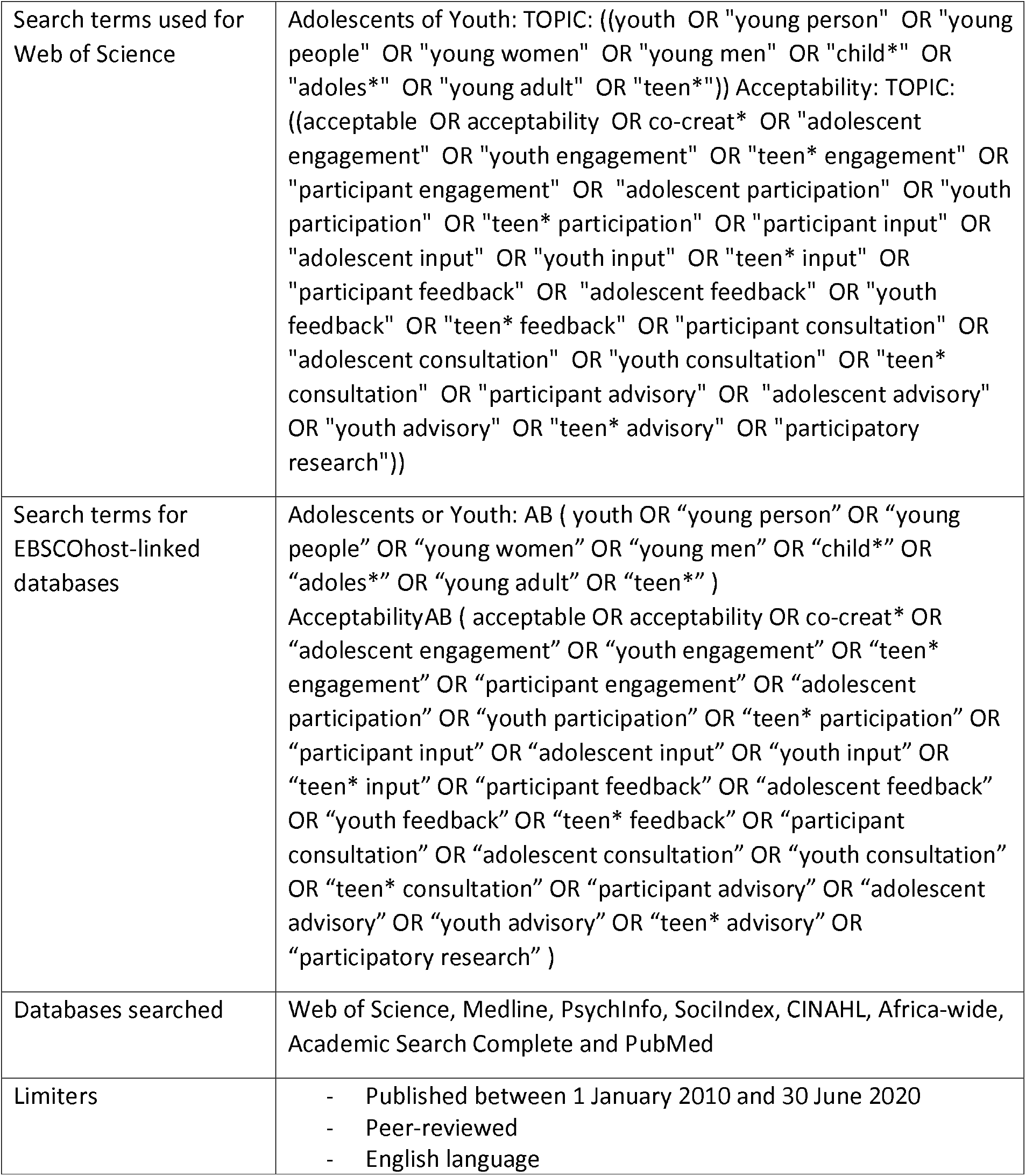
Systematic Review Search Strategy.

